# DAGM: a novel modelling framework to assess the risk of HER2-negative breast cancer based on germline rare coding mutations

**DOI:** 10.1101/2021.01.05.21249253

**Authors:** Mei Yang, Yanhui Fan, Zhi-Yong Wu, Jin Gu, Zhendong Feng, Qiangzu Zhang, Shunhua Han, Zhonghai Zhang, Xu Li, Yi-Ching Hsueh, Xiaoling Li, Jieqing Li, Meixia Hu, Weiping Li, Hongfei Gao, Ciqiu Yang, Chunming Zhang, Liulu Zhang, Teng Zhu, Minyi Cheng, Fei Ji, Juntao Xu, Hening Cui, Guangming Tan, Michael Q. Zhang, Changhong Liang, Zaiyi Liu, You-Qiang Song, Gang Niu, Kun Wang

**Affiliations:** Department of Breast Cancer, Cancer Centre, Guangdong Provincial People’s Hospital, Guangdong Academy of Medical Sciences, Guangzhou, Guangdong, China; Phil Rivers Technology, Beijing, China; Phil Rivers Technology, Shenzhen, China; Department of Oncology Surgery, Diagnosis and Treatment Centre of Breast Diseases, Shantou Affiliated Hospital, Sun Yat-sen University, Shantou, Guangdong, China; School of Biomedical Sciences, The University of Hong Kong, Hong Kong, China; Institute of Bioinformatics, University of Georgia, Athens, GA, USA; State Key Laboratory of Computer Architecture, Institute of Computing Technology, Chinese Academy of Sciences, Beijing, China; MOE Key Laboratory of Bioinformatics; Bioinformatics Division and Centre for Synthetic & Systems Biology, TNLIST; School of Medicine, Tsinghua University, Beijing, China; Department of Radiology, Guangdong Provincial People’s Hospital, Guangdong Academy of Medical Sciences, Guangzhou, Guangdong, China; BNRIST Bioinformatics Division, Department of Automation, Tsinghua University, Beijing, China

**Keywords:** HER2-negative Breast cancer, germline rare coding mutations, pathogenesis, immune suppression

## Abstract

**Background:** Breast cancers can be divided into HER2-negative and HER2-positive subtypes according to the status of *HER2* gene. Despite extensive studies connecting germline mutations with possible risk of HER2-negative breast cancer, the main category of breast cancer, it remains challenging to accurately assess its potential risk and to understand the potential mechanisms.

**Methods:** We developed a novel framework named Damage Assessment of Genomic Mutations (DAGM), which projects rare coding mutations and gene expressions into Activity Profiles of Signalling Pathways (APSPs).

**Findings:** We characterized and validated DAGM framework at multiple levels. Based on an input of germline rare coding mutations, we obtained the corresponding APSP spectrum to calculate the APSP risk score, which was capable of distinguish HER2-negative from HER2-positive cases. These findings were validated using breast cancer data from TCGA (AUC = 0.7). DAGM revealed the *HER2* signalling pathway was up-regulated in the germline of HER2-negative patients, and those with high APSP risk scores had suppressed immunity. These findings were validated using RNA sequencing, phosphoproteome analysis, and CyTOF. Moreover, using germline mutations, DAGM could evaluate the risk of developing HER2-negative breast cancer, not only in women carrying *BRCA1/2* mutations, but also in those without known disease-associated mutations.

**Interpretation:** The DAGM can facilitate the screening of subjects at high risk of HER2-negative breast cancer for primary prevention. This study also provides new insights into the potential mechanisms of developing HER2-negative breast cancer. The DAGM has the potential to be applied in the prevention, diagnosis, and treatment of HER2-negative breast cancer.

**Funding:** This work was supported by the National Key Research and Development Program of China (grant no. 2018YFC0910406 and 2018AAA0103302 to CZ); the National Natural Science Foundation of China (grant no. 81202076 and 82072939 to MY, 81871513 to KW); the Guangzhou Science and Technology Program key projects (grant no. 2014J2200007 to MY, 202002030236 to KW); the National Key R&D Program of China (grant no. 2017YFC1309100 to CL); and the Natural Science Foundation of Guangdong Province (grant no. 2017A030313882 to KW)

**Research in context:** *Evidence before this study:* The majority of hereditary breast cancers are caused by BRCA1/2 mutations, and the presence of these mutations is strongly associated with an increased risk of breast cancer. Meanwhile, BRCA1/2 gene mutations are rarely found in sporadic breast cancers and only account for a modest percentage of all breast cancer patients. Polygenic risk score (PRS), a widely-used approach for stratifying individuals according to their risk of a certain kind of complex disease, has been used to predict subjects at high risk for breast cancer. However, relying on SNPs from genome-wide association studies (GWAS) without including gene expressions or pathway activities, PRS is not very suitable for cross-population prediction and describes disease risk in terms of genomic mutations without alluding to the underlying pathogenic mechanism(s). Therefore, there is still an urgent need for a population-independent comprehensive method to accurately assess the risk of breast cancer and to gain insights on potential mechanism(s).

*Added value of this study:* When subjecting germline rare coding mutations (gRCMs) to DAGM framework, which results in the corresponding APSP and APSP risk score. Both APSP and APSP risk score can identify HER2-negative from HER2-positive breast cancers. These findings suggest HER2-negative breast cancer does not develop accidentally, but rather is defined by a genomic evolutionary strategy. Furthermore, this study also revealed the up-regulation of HER2 signalling pathway in germlines of HER2-negative breast cancers and the immune suppression in subjects with high APSP risk score, shedding new light on the potential mechanisms of developing HER2-negative breast cancer. Moreover, our APSP risk score was able to relatively accurately evaluate the risk of developing HER2-negative breast cancer for each female, including not only BRCA1/2 carriers, but also non-carriers.

*Implications of all the available evidence:* The present study suggests that *HER2* signalling pathway activity, as an aggressive factor, contribute to the development of different types of breast cancers, either via the combined effects of multiple germline mutations in HER2-negative germlines or via amplifying the gene itself in HER2-positive tumour cells. This provides a theoretical basis for the prevention, diagnosis, and treatment of breast cancers. At the same time, the study provides preliminary methods for assessing the relative risk of HER2-negative breast cancer for females with or without BRCA1/2 mutations. Finally, our findings provide a new perspective and theoretical basis for identifying high-risk female subjects, based on the high APSP risk score, for early screening and prevention of HER2-negative breast cancer.

## 1. Introduction

Breast cancer is the most common malignancy and the second leading cause of cancer death in women worldwide [1,2]. Breast cancer can be classified into two types: human epidermal growth factor 2 (HER2)-positive and HER2-negative, in which the former is characterized by HER2 amplification in tumour tissues, whereas the latter is not. The majority (80%) of breast cancers cases are HER2-negative, including triple-negative breast cancer (TNBC), luminal A, and luminal B (HER2-). Although germline mutations in genes such as *BRCA1/2* are known to be associated with breast cancer, they are detected in only 5.3% of breast cancer patients and 11.2% of TNBC patients in China [3]. Signalling pathways such as STING pathway and immunosuppressive pathways have been linked to the development and progression of breast cancers [4-6]. However, it still remains elusive to accurately assess the risk and understand the underline mechanisms of developing sporadic HER2-negative breast cancer.

To address these issues, extensive studies have been carried out in the past decade, leading to various methods such as Gene Set Enrichment Analysis (GSEA) [7] and Polygenic risk score (PRS) [8]. The PRS has been widely used to stratify individuals according to their risk for complex diseases including breast cancer [9-11]. The PRS relies on SNPs from genome-wide association studies (GWAS) without including gene expressions or pathway activities. Thus, it does not allude to the underlying pathogenic mechanism but describes the risk in terms of the association between genomic mutations and the disease. In predicting breast cancer risk, the performance of PRS models demonstrated an area under receiver-operator curve (AUC) of around 0.6 [9-12]. Due to the ancestry-dependence of GWAS, PRS is only suitable for risk stratification of individuals from the same cohort rather than across populations. Therefore, there is an urgent need for a population-independent comprehensive method, which can integrate genome mutations, gene expressions, and pathway activities to accurately assess the risk of breast cancer and to gain insights on the pathogenesis.

To meet these challenges, we developed a new framework, called Damage Assessment of Genomic Mutations (DAGM), which integrates genome-wide information of rare coding mutations and gene expression from the COSMIC cell line project to calculate the activity profile of signalling pathways (APSPs). When subjecting germline mutations to DAGM, we obtained the corresponding APSP spectrum. Using this APSP spectrum for HER2-negative breast cancer germlines, we found that the *HER2* signalling pathway was up-regulated compared to that in HER2-positive germlines, which was validated by RNA sequencing (RNA-Seq) and phosphoproteome analysis. Next, we calculated the germline APSP risk score and found it was able to distinguish between HER2-negative and HER2-positive breast cancers (AUC=0.79). This result was validated using the TCGA breast cancer cohort (AUC=0.70). Furthermore, subjects with high APSP risk scores demonstrated enhanced immune suppression, as determined by single-cell mass cytometry (CyTOF) analysis. Moreover, the germline APSP risk score could be used to evaluate the relative risk of HER2-negative breast cancer for each individual. These results show that DAGM provides relatively accurate assessment of HER2-negative breast cancer risk based on genome-wide comprehensive data of rare coding mutations and gene expression, which can facilitate high-risk subject screening, and also sheds new light on the potential pathogenic disease mechanisms

## 2. Materials and Methods

### 2.1 Overview

To accurately assess the risk of breast cancer and to understand the underlying mechanisms and pathogenesis, we developed the new DAGM framework to mine genomic information including rare coding mutations, gene expressions, and signalling pathways activities. Instead of looking for specific genes that determine the specific fate of cells, we assume the unique fate of a cell is determined by a set of genes through regulating the activities of functional modules such as signalling pathways. First, we constructed a framework based on public data from the COSMIC cell line project. After validating its reliability, we then applied the framework to whole-exome data from breast cancer patients and controls. The results showed our framework could identify subjects at high risk of HER2-negative breast cancer and also provided clues for the potential mechanisms underlying the disease pathogenesis. We validated our findings by RNA-Seq, phosphoproteome analysis, and CyTOF analysis, as well as using breast cancer data from TCGA.

### 2.2 Study participants

This study recruited 721 subjects including 434 breast cancer patients and 287 controls. The average age of the controls was 81 years. Patients with breast cancer were recruited from two independently operated hospitals in Guangdong province in southern China (Table 1), including 316 patients in the Guangdong Provincial People’s Hospital (GZ cohort: 62 ERBB2-positive and 43 Luminal B (HER2+), 77 Luminal B (HER2-), 24 Luminal A, and 110 TNBC) and 118 patients from the Shantou Affiliated Hospital (ST cohort: 15 ERBB2-positive and 103 TNBC). A total of 287 cancer-free Chinese females from Hong Kong including ApoE LJ4-negative late-onset Alzheimer’s disease cases and age-matched controls [13] were used as the controls.

**Table 1.** Study population characteristics.

| **Characteristic** | **Luminal A** | **Luminal B**<br>**HER2-** | **Luminal B**<br>**HER2+** | **ERBB2-**<br>**positive** | **TNBC** |
| --- | --- | --- | --- | --- | --- |
| **Age, Mean (range , years)** | 50(38-71) | 47(22-80) | 48(25-63) | 51(22-72) | 50(23-82) |
| **Histological grade, n (%)** |  |  |  |  |  |
| **1** | 1/24 (4.2) | 1/77(1.3) | 0/43 (0) | 0/77 (0) | 5/213 (2.3) |
| **2** | 14/24 (58.3) | 36/77(46.8) | 23/43(53.5) | 28/77(36.4) | 78/213(36.6) |
| **3** | 7/24 (29.2) | 31/77(40.3) | 16/43(37.2) | 40/77(51.9) | 109/213(51.2) |
| **Unknown** | 2/24 (8.3) | 9/77 (11.7) | 4/43(9.3) | 9/77(11.7) | 21/213(9.9) |
| **Tumour size, n (%)** |  |  |  |  |  |
| **≤2cm** | 6/24(25.0) | 24/77(31.2) | 10/43(23.3) | 11/77(14.3) | 51/213(23.9) |
| **> 2cm** | 15/24 (62.5) | 50/77 (64.9) | 31/43(72.1) | 57/77(74.0) | 157/213(73.7) |
| **Unknown** | 3/24 (12.5) | 3/77 (3.9) | 2/43(4.7) | 9/77(11.7) | 5/213(2.3) |
| **Nodal status, n (%)** |  |  |  |  |  |
| **LN-** | 13/24 (54.2) | 37/77 (48.1) | 24/43(55.8) | 31/77(40.3) | 126/213(59.2) |
| **LN+** | 8/24(33.3) | 36/77(46.8) | 17/43(39.5) | 38/77(49.4) | 82/213(38.5) |
| **Unknown** | 3/24 (12.5) | 4/77(5.2) | 2/43(4.7) | 8/77(10.4) | 5/213(2.3) |
| **Ki 67 status, n (%)** |  |  |  |  |  |
| **≤10** | 2/24 (8.3) | 11/77(14.3) | 8/43(18.6) | 5/77(6.5) | 20/213(9.4) |
| **10-30** | 7/24 (29.2) | 23/77(29.9) | 13/43(30.2) | 23/77(29.9) | 47/213(22.1) |
| **> 30** | 14/24 (58.3) | 40/77(51.9) | 21/43(48.8) | 48/77(62.3) | 136/213(63.8) |
| **unknown** | 1/24 (4.2) | 3/77(3.9) | 1/43(2.3) | 1/77(1.3) | 10/213(4.7) |

### 2.3 Ethics

The study protocol was approved by the Research Ethics Committee at Guangdong General Hospital, Guangdong Academy of Medicals Sciences. Written informed consent was obtained from all participants for the use of banked tissue (including white blood cells and buccal cells) and for the collection of pathological data and clinical follow-up data.

### 2.4 Whole-exome sequencing and variant calling

Peripheral blood mononuclear cell (PBMC) samples were collected from breast cancer patients. DNA was extracted using QIAamp DNA mini kit (Qiagen) according to the blood and body fluid protocol in the user manual. Paired-end multiplex sequencing of case samples was performed on an Illumina HiSeq X Ten sequencing platform to a median depth of 150-250X. Samples from the control cohort were sequenced on an Illumina HiSeq 2000 system to a median depth >60X. Paired-end raw sequence reads were mapped to the human reference genome (UCSC hg19) using the Burrows-Wheeler Aligner [14] with default settings. Variant calling was carried out using the Genome Analysis Toolkit (GATK) with the HaplotypeCaller module [15] according to GATK Best Practices. Briefly, the aligned BAM files were first marked for duplicate reads by Picard. Local realignment around indels and base quality score recalibration were performed using GATK. The processed BAM files were then used to call SNPs and indels. Variant filtering of SNPs and indels were performed separately by variant quality score recalibration using GATK. The filtered variants were then annotated by ANNOVAR [16].

### 2.5 Transcriptomics

A total of 3 µg of RNA per sample was used as the input material for RNA sample preparation. Sequencing libraries were generated using NEBNext® UltraTM RNA Library Prep Kit for Illumina® (NEB, USA) following the manufacturer’s instructions. Index codes were added to attribute the sequences to each sample. Sequencing was performed on an Illumina HiSeq platform. Raw data (raw reads) in FASTQ format were processed through our in-house Perl scripts to clean the data (clean reads), which removed reads containing adapters, reads containing poly-N, and low quality reads. All downstream analyses were based on high-quality clean data. Paired-end clean reads were aligned to the reference genome using Hisat2 v2.2.1 [17]. FeatureCounts v2.0.1 [18] was used to count the numbers of reads mapped to each gene. Only genes with at least one count per million in at least two samples were kept for the following analysis. Read counts were normalized using the TMM normalization method performed in the edgeR package v3.26.8 [19]. The RPKM values of the genes were calculated based on the length of the gene and read counts mapped to the gene.

### 2.6 Phosphoproteome analysis

Samples were minced individually in liquid nitrogen. The enrichment was carried out using PHOS-Select iron affinity gel (Sigma, P9740) following the manufacturer’s instructions. Shotgun proteomics analyses were performed using an EASY-nLC™ 1200 UHPLC system (Thermo Fisher) coupled with an Orbitrap Q Exactive HF-X mass spectrometer (Thermo Fisher) operating in a data-dependent acquisition (DDA) mode. The resulting spectra from each fraction were searched separately against the UniProt database [20]. For protein identification, proteins with at least one unique peptide were identified at an FDR less than 1.0% at the peptide and protein level, respectively.

### 2.7 CyTOF

Antibodies were either purchased pre-conjugated (Fluidigm, DVS Sciences) or purchased and conjugated in-house using MaxPar X8 Polymer Kits (Fluidigm) according to the manufacturer’s instructions (Suppl. Table 1). Scans were acquired on a Helios 2.0 (Fluidigm) at an event rate of 300 events/s. After normalizing and randomizing values to near zero using the Helios software, FCS files were then generated for the analysis. Mass cytometry data was de-barcoded using a doublet filtering scheme with mass-tagged barcodes, and then manually gated to retain live, singlet, and valid immune cells. Data generated from different batches were normalized through the bead normalization method. All cell events in each individual sample were pooled and included in the analysis.

### 2.8 The Damage Assessment of Genomic Mutations (DAGM) framework

Similar to the eQTL method that uses gene expression as an independent trait to determine its linkage site in the genome, we can correlate the gene expression in cell lines to ‘traits’ and associate them with a variety of different genes with rare coding mutations (RCMs). However, rather than examining how all genes with rare mutations change the gene expression traits, DAGM screens out genes defined as global driver genes, whose mutations deterministically cause almost the same alterations in the global gene expression across different cellular or tissue contexts. For example, the well-known driver gene *TP53* drives similar altered gene expressions in tumour cells from different origins. We are continuously collecting RCM data and gene expression traits from cancer cell lines or primary cells to improve the database and to provide more accurate global driver genes. Based on a sample with only RCM data, DAGM was able to determine candidate genes based on the RCM distribution, in which the effects of driver genes were combined to give the altered gene expression, representing the mutations as an overall impact on gene expression. The DAGM can then determine the activity profiles of signalling pathways (APSP) in three sequential steps (see supplementary method for full details). The first step takes the mutated genes and gene expressions as the input and calculates the effect of the rare mutations on gene expression changes within the cell lines, which will generate a *n x n* matrix. Where each row represents a mutated gene, and each column denotes the expression of a gene, and the value in the cell was defined a driving force and the global driving force (GDF) is calculated to select global driving genes. The second step is to calculate the combined effect of all the mutations an individual carries. If an individual carries *m* mutated genes, this step converts the *m x n* matrices of different dimensions into *1 x n* matrices in the same dimension, which can be compared between different samples. The third step is to evaluate the activity profiles of signalling pathways (APSPs). In this study, we included 60 pathways (Table 2), so DAGM will output a *1 x 60* matrix for each sample.

**Table 2.** Signalling pathways for DAGM framework.

| **Index** | **Pathway** | **Pathway Panel** |
| --- | --- | --- |
| 1 | CDK5 | Development |
| 2 | Hedgehog | Development |
| 3 | NOTCH | Development |
| 4 | Hypoxia | Response to stress |
| 5 | ROS | Response to stress |
| 6 | Unfolded protein response | Response to stress |
| 7 | UVB induced MAPK | Response to stress |
| 8 | UV response | Response to stress |
| 9 | Adipogenesis | Energy & metabolism |
| 10 | AMPK | Energy & metabolism |
| 11 | mTORC1 | Energy & metabolism |
| 12 | mTOR | Energy & metabolism |
| 13 | Beta Cells | Energy & metabolism |
| 14 | PI3K-AKT-mTOR | Energy & metabolism |
| 15 | PI3K-AKT | Energy & metabolism |
| 16 | PPARA-RXRA | Energy & metabolism |
| 17 | PKA | Energy & metabolism |
| 18 | VDR-RXR | Energy & metabolism |
| 19 | 4-1BB | Immune system |
| 20 | BCR | Immune system |
| 21 | CD40 | Immune system |
| 22 | IL2-Stat5 | Immune system |
| 23 | INF alpha | Immune system |
| 24 | INF gamma | Immune system |
| 25 | Jak-Stat | Immune system |
| 26 | LBC | Immune system |
| 27 | Nf-kappaB | Immune system |
| 28 | Toll like receptor | Immune system |
| 29 | AML | Pro-tumour |
| 30 | Angiogenesis | Pro-tumour |
| 31 | Angiopoietin | Pro-tumour |
| 32 | Colorectal cancer Metastasis | Pro-tumour |
| 33 | Estrogen-dependent BRCA | Pro-tumour |
| 34 | Glioblastoma | Pro-tumour |
| 35 | Glioma invasiveness | Pro-tumour |
| 36 | Glioma | Pro-tumour |
| 37 | NSCLC | Pro-tumour |
| 38 | Pancreatic adenocarcinoma | Pro-tumour |
| 39 | Renal cell carcinoma | Pro-tumour |
| 40 | Renin angiotensin | Pro-tumour |
| 41 | Apoptosis | Anti-tumour |
| 42 | Ceramide | Anti-tumour |
| 43 | TP53 | Anti-tumour |
| 44 | PTEN | Anti-tumour |
| 45 | DNA synthetic checkpoint | Cell cycle & proliferation |
| 46 | DNA damage checkpoint | Cell cycle & proliferation |
| 47 | Erk/Mapk | Cell cycle & proliferation |
| 48 | KRAS | Cell cycle & proliferation |
| 49 | Telomerase | Cell cycle & proliferation |
| 50 | EGF | Growth factors |
| 51 | Her2 | Growth factors |
| 52 | ErbB4 | Growth factors |
| 53 | FGF | Growth factors |
| 54 | HGF | Growth factors |
| 55 | IGF-1 | Growth factors |
| 56 | PDGF | Growth factors |
| 57 | TGF-beta | Growth factors |
| 58 | VEGF | Growth factors |
| 59 | Wnt beta-catenin | Growth factors |
| 60 | Wnt calcium | Growth factors |

### 2.9 Classification

Binary classifiers were built using the support vector machine (SVM) and logistic regression implemented in the caret R package. All data were randomly split into the training dataset (60% or 75%) and the testing dataset (40% or 25%), while preserving the overall class distribution of the data. The optimal parameters were selected by five-fold cross validation. The criteria of the area under the receiver operating characteristics curve (AUC) was used to measure the performance of the classifiers.

### 2.10 Activity profiles of signalling pathway (APSP) risk score

Three pathway panels including growth factor, pro-tumour, and immune function pathways were significantly downregulated in HER2-negative breast cancers and were selected to calculate the APSP risk score. The APSP risk score was calculated as the weighted mean of the APSPs of the three selected pathway panels in each person. The weights were *-1/n* and *1/n* for the *HER2* signalling pathway and for other pathways, respectively.

### 2.11 Statistical analysis

The pheatmap R package was used to plot the clustered heatmaps using Ward’s criteria [21]. The ggpubr R package was used to draw the scatter plots, boxplots, bar plots, histograms, and linear regression lines with 95% confidence interval. The plotROC R package was used to draw ROC. Pearson correlation coefficients (PCC) and P-values were also labelled on the plots. The difference between two groups was analysed by Student’s t test. A P-value < 0.05 was considered statistically significant. All statistical analyses and plots were conducted using in-house scripts developed in Python, Perl, and R.

### 2.12 Availability of data and material

The raw exome sequencing data reported in this paper have been deposited in the Genome Sequence Archive (GSA) in National Genomics Data Centre, Beijing Institute of Genomics, Chinese Academy of Sciences, under accession number HRA000285, which is publicly accessible at https://bigd.big.ac.cn/gsa. All other data and materials are available upon request.

## 3 Results

### 3.1 Characterization and validation of the DAGM framework

We developed a novel framework called Damage Assessment of Genomic Mutations (DAGM), which consists of three sequential steps (Fig. 1, see supplementary method for details). The first step takes the mutated genes and gene expressions as the inputs to determine the driving force of each mutated gene on the expression of all genes as the output. The global driving force (GDF) is then calculated and the global driving genes are identified. The second step calculates the combined effects of all mutations that an individual carries. The third step evaluates the activity profiles of signalling pathways (APSPs).

**Fig. 1.**
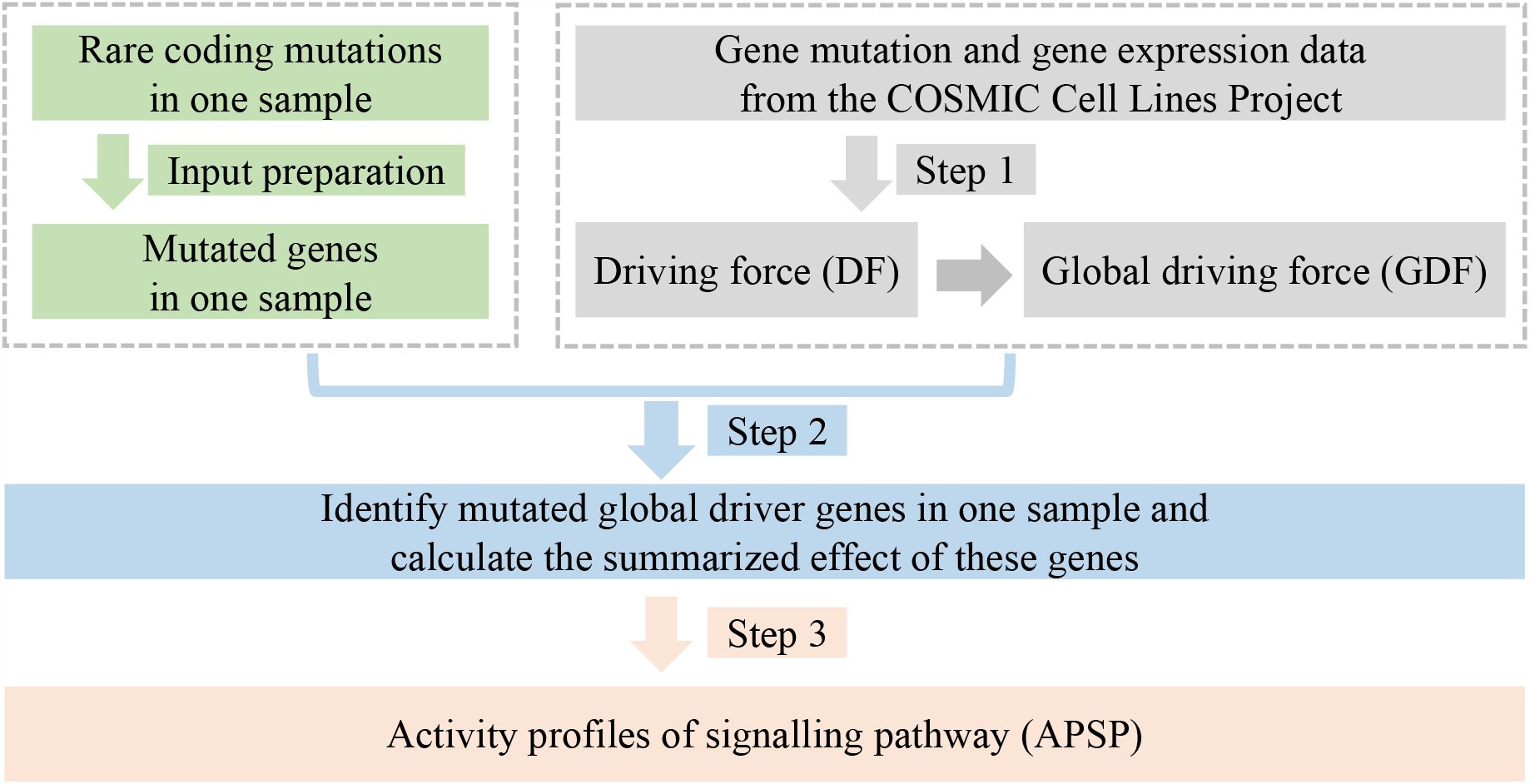
Workflow of the DAGM framework. The DAGM framework mainly composed of three steps. In the first step (grey colour), driving force (DF) and global driving force (GDF) are established for future analysis based on the rare coding mutations and gene expression from the COSMIC Cell Lines project. When obtaining the germline rare coding mutations of a subject, we calculated the combined effect of all these mutations (Step2). In the last step, the activity profile of signalling pathways (APSPs) is evaluated for this subject.

To identify the global driver genes regardless of different extrinsic/intrinsic cellular conditions, we assessed the driving force based on the genome-wide data of rare coding mutations and gene expression in 970 cancer cell lines (COSMIC Cell Line project), considering cancer cell lines exhibit high tumour purity and low expression heterogeneity. To validate this framework, we used another dataset from Cancer Cell Line Encyclopaedia (CCLE) [22] to build a ‘driving force’ matrix. The GDFs calculated from the two different datasets were significantly correlated (coefficient of determination: R2 = 0.99, P-value < 1.0×10-16, Suppl. Fig. 1a), suggesting the first step of DAGM is robust across different datasets. Next, we selected the top 10 genes with highest GDF in the DAGM and searched public databases of cancer driver genes. We found six of these genes, *KRAS, TP53, MYC, BCL2, BRAF*, and *RPL22*, were listed in COSMIC Cancer Gene Census [23], whereas all 10 were listed in DriverDBv3 [24], which confirmed the potential ability of using GDF to identify essential genes for tumourigenesis, including cancer driver genes.

To test whether the APSP calculated from DAGM reflects signalling pathway activities, we applied DAGM to somatic mutations from tumour samples from 16 ERBB2-positive patients and 42 TNBC patients (Suppl. Fig. 1b). Interestingly, although HER2 amplification was not included in the input mutation information, the resultant APSP showed significantly higher *HER2* signalling pathway activity (T-test, P-value = 0.00095) in ERBB2-positive breast tumours compared to TNBC (Suppl. Fig. 1c). This result was consistent with the upregulation of the *HER2* signalling pathway in HER2-positive breast cancer samples, which was also demonstrated by fluorescence in situ hybridization (FISH) [25].

### 3.2 APSP spectrum based on germline mutations distinguishes between HER2-negative and HER2-positive breast cancers

As is well known, it is of great importance to distinguish HER2-negative from HER2-positive breast cancer, but this currently relies on *HER2* FISH of tumour tissues. To test whether the APSP from DAGM can be used to identify breast cancer subtypes, we collected germline rare coding variants from 721 subjects, which included 434 breast cancer patients of different subtypes and 287 cancer-free female subjects in the control group. The DAGM analysis using these input germline mutations (Suppl. Fig. 2) revealed the APSP spectrums for HER2-negative patients were remarkably lower than that for HER2-positive patients or controls (Fig. 2a). Further hierarchical clustering using Euclidean distance showed that Luminal A, Luminal B (HER2-), and TNBC were grouped together, whereas ERBB2-positive and Luminal B (HER2+) were clustered into another group (Fig. 2b), suggesting the germline APSP spectrum can distinguish HER2-negative breast cancer from HER2-positive breast cancer subtypes. To quantitatively evaluate the performance of APSP in identifying breast cancer subtypes, we built a binary classifier based on the APSP of all 60 pathways using the support vector machine (SVM) with a radial basis function kernel with five-fold cross validation. The results showed an average value of the area under the receiver operating characteristic curve (AUC) of 0.77 for the classifiers between Her2-negative and Her2-positive patients (Fig. 2c), and 0.76 between Her2-negative and controls (Fig. 2d). Therefore, the APSP spectrum based on germline mutations can distinguish HER2-negative from HER2-positive breast cancer subtypes.

**Fig. 2.**
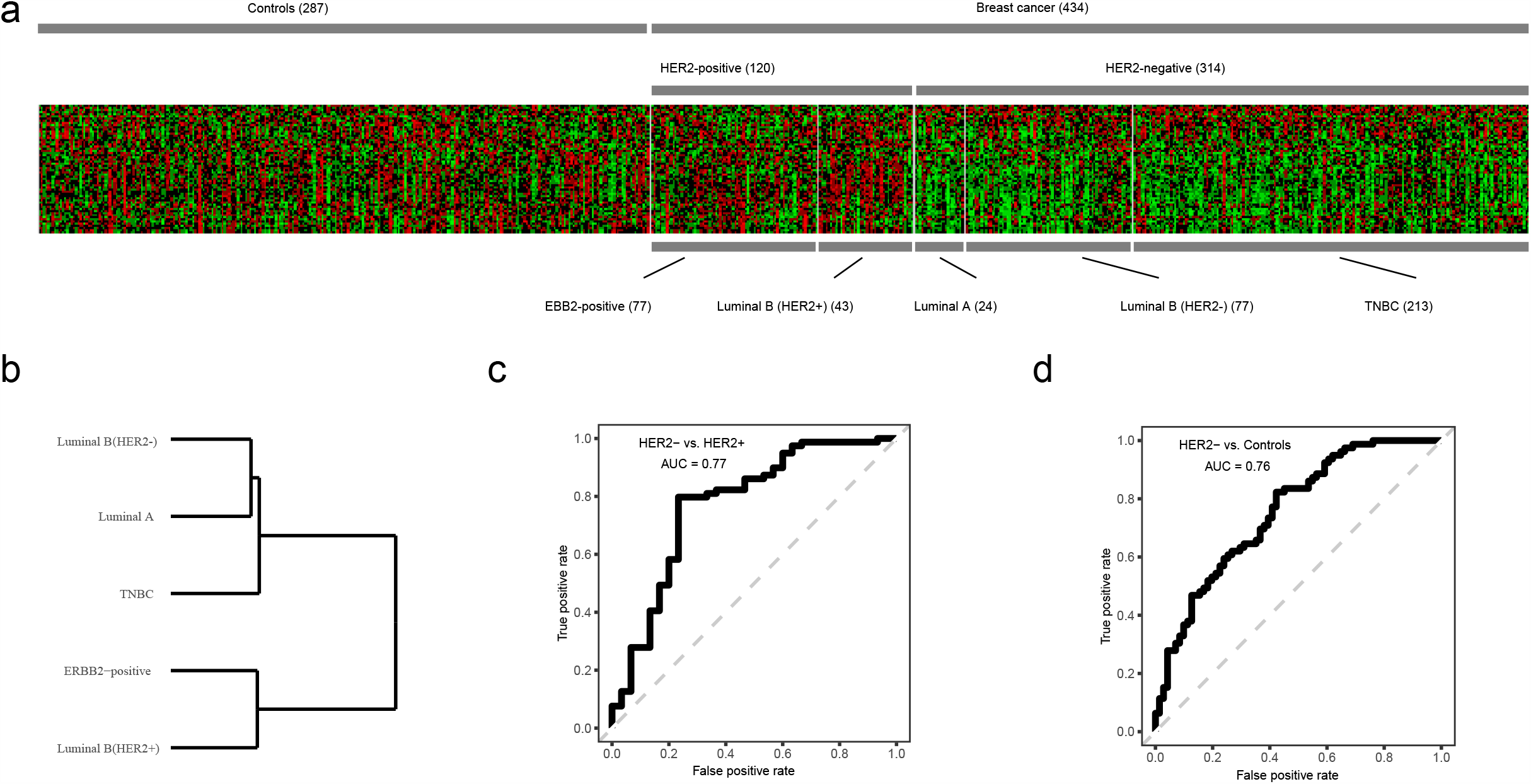
Germline APSP spectrum distinguishes HER2-negative breast cancer patients from HER2-positive patients. (a) Heatmap of the germline APSPs of 60 pathways for 721 subjects. Each row represents one pathway and each column represents one subject. (b) Hierarchical clustering dendrogram. HER2-negative subtype (Luminal B (HER2-), Luminal A, and TNBC) and HER2-positive subtype (ERBB2-positive and Luminal B (HER2+)) were well separated by hierarchical clustering with Euclidean distance as the distance measure. (c) Receiver operating characteristic (ROC) curve for distinguishing HER2-negative breast cancer patients from HER2-positive patients by APSPs. Area under the curve (AUC) was 0.77. (d) Receiver operating characteristic (ROC) curve for distinguishing HER2-negative breast cancer patients from controls by APSPs. Area under the curve (AUC) was 0.76.

### 3.3 APSP reveals up-regulation of the HER2 signalling pathway in the germlines of HER2-negative breast cancer

Although *BRCA1/2* mutations are currently used in breast cancer screening [26-28], carriers of these mutations represent only in a small number of HER2-negative breast cancer patients. To find out the potential signalling features that are characteristic of HER2-negative breast cancer, we further examined the APSPs in more depth. Interestingly, most cellular signalling pathways were drastically down-regulated in HER2-negative patients compared to control subjects, whereas almost no significant changes in HER2-positive patients were observed (Fig. 3a). Next, we applied TCGA data to crosscheck the activity differences in the 60 pathways between HER2-positive and HER2-negative patients, which showed significant correlations with our results (Pearson’s correlation coefficient = 0.93, P-value = 3.8e-27, Suppl. Fig. 3a). Furthermore, the Z-scores of the APSPs of 60 signalling pathways were strongly correlated between HER2-negative subtypes (Luminal B (HER2-) and TNBC) (Pearson’s correlation coefficient = 0.86, P-value = 1.14e-18, Fig. 3b) but not for HER2-positive subtypes (Luminal B (HER2+) vs. ERBB2-positive) (Pearson’s correlation coefficient = 0.56, P-value = 3.92e-6, Fig. 3c). These findings suggest that HER2-negative breast cancer can be characterized by germline APSP features.

**Fig. 3.**
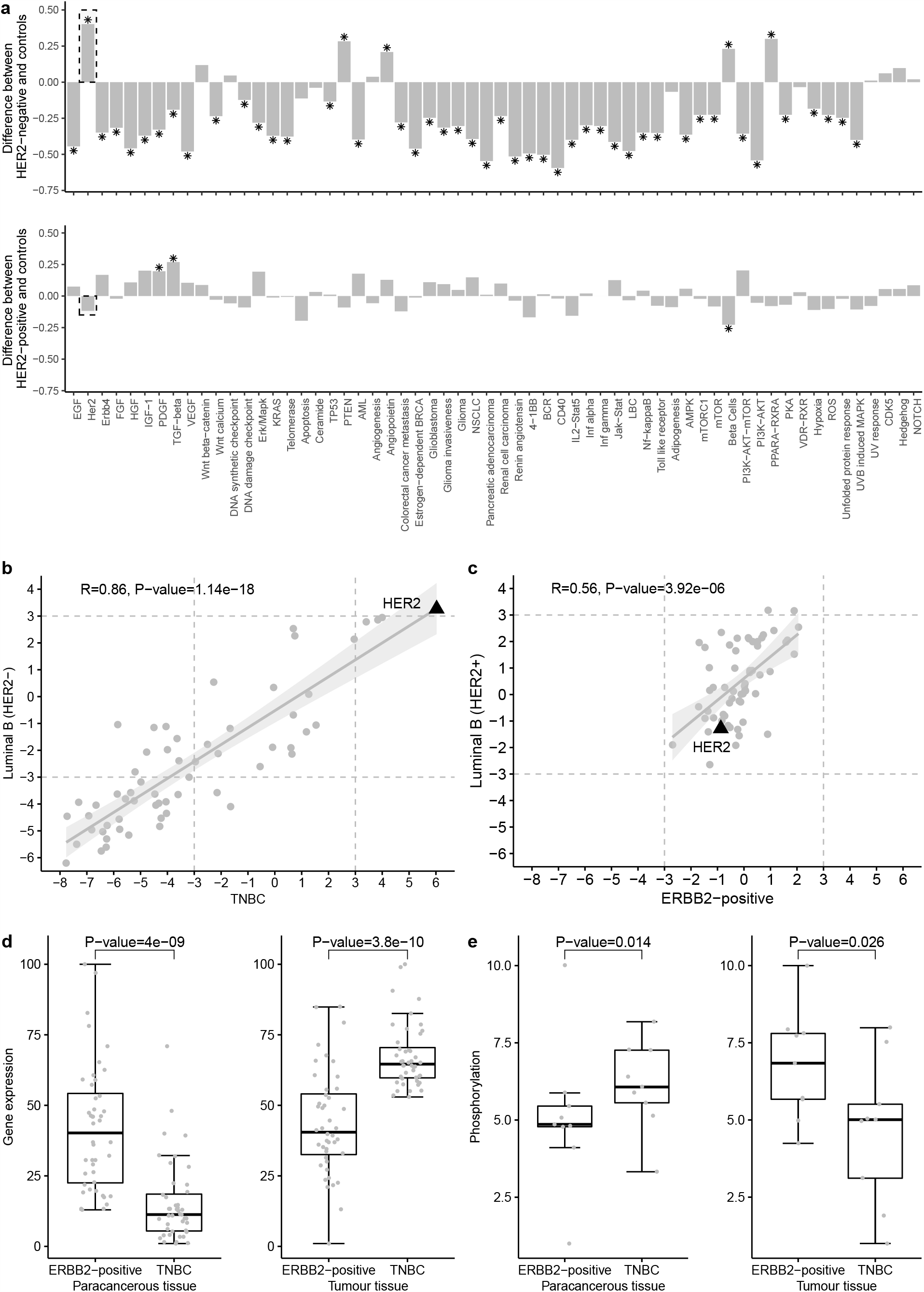
Upregulated HER2 signalling pathway in germlines of Her2-negative breast cancer patients. (a) Difference of activity profiles of signalling pathways (APSPs) between breast cancer patients and controls. To test the significance of the difference, P-values were calculated by permutation (1,000,000 times). The FDR corrected P-values were also calculated to correct the multiple comparisons. The asterisk denotes FDR corrected P-valueLJ<LJ0.05. Most pathways activities were significantly down-regulated in HER2-negative breast cancer germlines (upper panel), whereas the HER2 signalling pathway activity was significantly up-regulated (rectangle, upper panel). However, only a few pathway activities were significantly altered in HER2-positive breast cancer patients (lower panel). (b) Correlation of APSPs between TNBC and Luminal B (HER2-). For HER2-negative breast cancer, germline pathway APSPs between TNBC and Luminal B (HER2-) subtype were strongly positively correlated. The black triangle represents the HER2 signalling pathway. (c) Correlation of APSP between ERBB2-positive and Luminal B (HER2+). For HER2-positive breast cancer, germline pathway APSPs between ERBB2-positive and Luminal B (HER2+) subtype were weakly positively correlated. (d) RNA-Seq validation of the upregulated HER2 signalling pathway. We performed RNA-Seq on both cancerous and paracancerous tissues from TNBC and ERBB2-positive breast cancer patients, the expression level of HER2 pathway signature genes in paracancerous tissue were significantly lower in the TNBC compared to ERBB2-positive patients, which validated the upregulated HER2 signalling pathway in HER2-negative breast cancer germlines. (e) The phosphoproteome validation the upregulated of the HER2 signalling pathway. Phosphoproteome analysis of both cancerous and paracancerous tissues from TNBC and ERBB2-positive breast cancer patients validated the upregulated HER2 signalling pathway in HER2-negative breast cancer germlines.

Among the significantly altered pathways according to APSP, we found significant up-regulation of the *HER2* signalling pathway in germlines of HER2-negative breast cancer patients, even though *HER2* gene was not amplified in the tumour tissues. To validate this finding, we performed RNA-Seq and phosphoproteome analysis using breast cancer and paracancerous (normal breast) tissues from both ERBB2-positive and TNBC patients. The expression of signature genes of the *HER2* signalling pathway in paracancerous tissues were systematically lower in TNBC patients than in ERBB2-positive patients (Fig. 3d, left panel), despite high expression of these genes in TNBC tumour tissues (Fig. 3d, right panel). These observations indicated upregulation of the *HER2* signalling pathway in TNBC paracancerous tissues and supported the conclusion inferred from APSP. Furthermore, the results from phosphoproteome analysis confirmed activation of the *HER2* signalling pathway in paracancerous tissues of TNBC compared to ERBB2-positive patients (Fig. 3e). Therefore, the upregulation of *HER2* signalling pathway in germlines revealed by APSP are a specific feature of HER2-negative patients, as validated by RNA-Seq and phosphoproteome data.

### 3.4 APSP risk score based on germline mutations is able to identify HER2-negative breast cancer

We demonstrated APSP spectrum could distinguish HER2-negative breast cancer from controls and HER2-positive breast cancer. Next, we classified the APSPs of the 60 signalling pathways into eight panels according to their functions. The pathways associated with the immune system, growth factor and pro-tumour pathways were significantly down-regulated in HER2-negative breast cancers (Fig. 4a), indicating these three panels are characteristic of HER2-negative breast cancer. Based on these three panels, we calculated the APSP risk score for each individual and found the risk scores were significantly higher in HER2-negative breast cancer patients than in controls (T-test, P-value = 3.96e-27) or in HER2-positive breast cancer patients (T-test, P-value = 4.88e-22) (Fig. 4b). A similar difference in the APSP risk scores was observed using the TCGA data (P-value = 5.4e-11, Suppl. Fig. 3b), suggesting the APSP risk score can be used to identify HER2-negative breast cancer. Given that women carrying *BRCA1/2* mutations have significantly increased risk for breast cancer [26-28], we compared the APSP risk scores for HER2-negative patients with or without *BRCA1/2* mutations. We found all HER2-negative patients had higher APSP risk scores, than control subjects regardless of *BRCA1/2* mutations (Fig. 4c T-test, P-value=9.5e-8 and P-value=3.1e-24 for *BRCA1/2* carriers and non-carriers, respectively). These findings imply that APSP risk score can be used to recognize not only HER2-negative patients with *BRCA1/2* mutations, but also those without any known biomarkers.

**Fig. 4.**
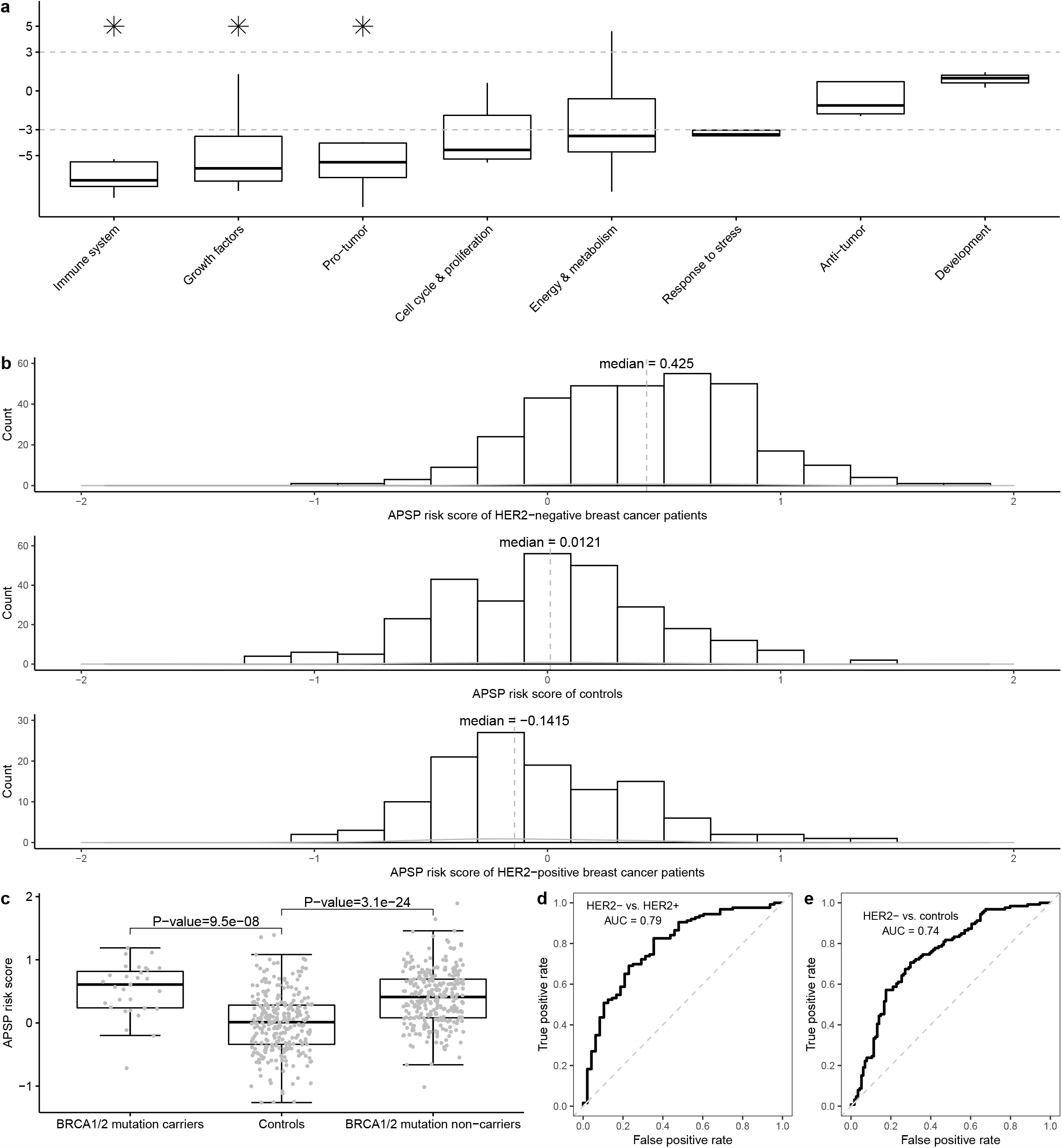
Identification of HER2-negative breast cancer using germline APSP risk score. (a) The boxplot of APSPs for eight pathway panels. The panel of immune system is at the top of three significantly down-regulated panels (labelled by a star) that were then used to calculate APSP risk scores. (b) The distribution of APSP risk scores of HER2-negative breast cancer patients (upper panel), controls (middle panel) and HER2-positive breast cancer patients (bottom panel). The APSP risk scores in HER2-negative breast cancer patients were significantly higher than those in the controls (T-test, P-value = 3.96e-27) and HER2-positive breast cancer patients (T-test, P-value = 4.88e-22). (c) The APSP risk scores were significantly higher in HER2-negative breast cancer patients with or without BRCA1/2 mutations than the control subjects. (d) Receiver operating characteristic (ROC) curve for APSP risk score in distinguishing HER2-negative breast cancer patients from HER2-positive patients. Area under the curve (AUC) was 0.79. (e) Receiver operating characteristic (ROC) curve for APSP risk score in distinguishing HER2-negative breast cancer patients from controls. Area under the curve (AUC) was 0.74.

To quantitatively evaluate the use of APSP risk score in identifying HER2-negative breast cancer, we built a binary classifier based on the APSP risk score using the logistic regression with five-fold cross validation. The results showed an AUC of 0.79 for the classifiers between HER2-negative and HER2-positive cases (Fig. 4d), and 0.74 for the classifier between HER2-negative and controls (Fig. 4e). Using data from TCGA, the logistic regression binary classifier could distinguish between HER2-negative and HER2-positive breast cancer patients with an average AUC value of 0.70 (Suppl. Fig. 3c). Taken together, germline APSP risk score can be used to identify HER2-negative breast cancer patients.

### 3.5 Enhanced immune suppression in subjects with high APSP risk scores

The APSP risk score was determined using three significantly downregulated pathway panels, including the immune system panel. Therefore, we reasoned that subjects with higher APSP risk scores would exhibit different altered immune states. To this end, we performed single-cell mass cytometry to count the number of different types of T cell types. Strikingly, we observed significantly increased exhausted CD8+ T lymphocytes in those with a high APSP risk score compared to those with a low risk score group (Fig. 5a, T-test, P-value = 0.0028), indicating an association between APSP risk score and immune suppression. Consistently, we also found significantly increased levels of exhausted CD8+ T lymphocytes in TNBC patients compared to controls (Fig. 5b, T-test, P-value = 0.00038), indicating enhanced immune suppression in TNBC patients with high APSP risk scores. These results revealed that subjects with higher APSP risk score also have suppressed immunity, which possibly contributes to an increased risk of HER2-negative breast cancer.

**Fig. 5.**
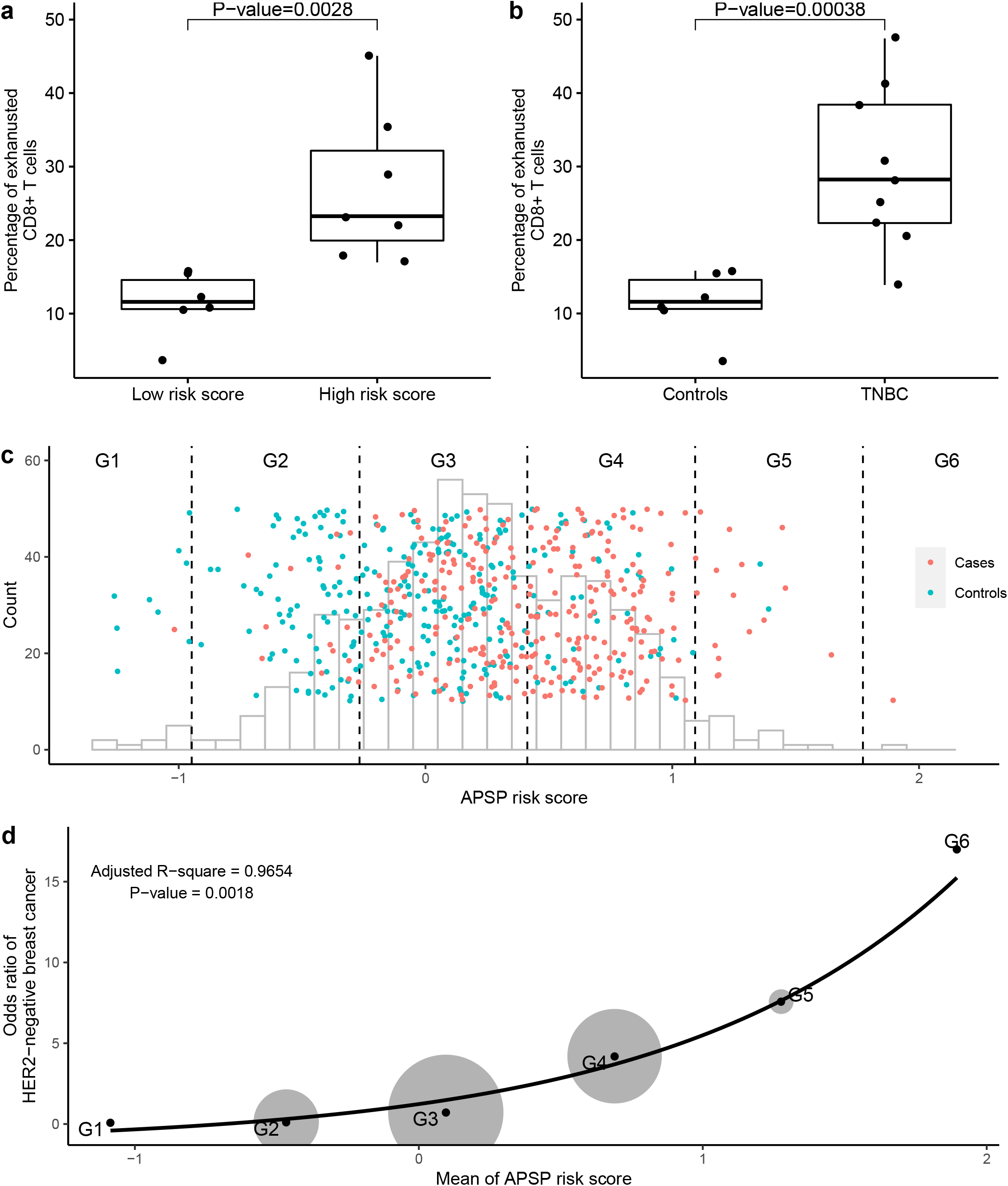
Enhanced immune suppression and risk stratification of HER2-negative breast cancer. (a) Mass cytometry analysis revealed percentage of exhausted CD8+ T cells in female subjects of low and high APSP risk scores. (b) Mass cytometry analysis revealed percentage of exhausted CD8+ T cells in TNBC patients and control subjects. (c) Stratification of HER2-negative patients (red dots) as well as control subjects (green dots) into six groups, G1 to G6, according to the different standard deviations of APSP risk scores from their mean value. G1 and G6 included subject with APSP risk score two standard deviations lower and higher than the mean value, respectively. G2 and G5 included subject with APSP risk score between one and two standard deviations lower and higher than the mean value, respectively. G3 and G4 included subject with APSP risk score within one standard deviation lower and higher than the mean value, respectively. (d) Odds ratios for HER2 negative breast cancer display exponential distribution when the subjects were stratified into six groups (G1 to G6) as in (c). The grey circles represent the sample sizes of each group.

### 3.6 APSP risk score based on germline mutations stratifies risk of HER2-negative breast cancer

As the APSP risk score could identify HER2-negative breast cancer subtype, we wondered if it could be used to quantitatively used to evaluate the risk of HER2-negative breast cancer. We divided the subjects into six groups (G1 to G6) and calculated their corresponding odds ratio for HER2-negative breast cancer. The odds ratio increased with increasing APSP risk score from G1 to G6 (Fig. 5c) and followed exponential distributions (Fig. 5d). There was only one patient in the highest risk group G6 and no controls. The odds ratio in group G5 reached 7.7, indicating these individuals have a high risk for HER2-negative breast cancer, whereas the odds ratio in G1 was 0.08, indicating these individuals have a much lower risk for HER2-negative breast cancer.

In summary, DAGM can identify HER2-negative breast cancer based on germline mutations and the calculated APSP risk score can be used to stratify the risk.

## 4. Discussion

HER2-negative breast cancer is the main category of breast cancer, the most common malignancy worldwide and the second leading cause of cancer death in women. It remains a challenge to evaluate the risk for developing HER2-negative breast cancer, despite extensive studies [9-12,29]. We developed a new framework, DAGM (Damage Assessment of Genomic Mutations). Different to other approaches [7,9-12,29,30], DAGM integrates gene mutations and gene expressions data to assess the risk and potential pathogenesis of a disease. DAGM can use this information to calculate the activity profiles of signalling pathways (APSPs) and then derive the APSP risk score to assess the disease risk in an individual subject. Using germline mutations, DAGM was able to distinguish HER2-negative breast cancer patients from HER2-positive patients. The derived germline APSP risk score was able to predict the risk of developing HER2-negative breast cancer, not only in those with BRCA1/2 mutations, but also in those without any known disease-associated mutations. Furthermore, DAGM revealed that the *HER2* signalling pathway was upregulated in the germline of HER2-negative patients, and those with high APSP risk scores also had enhanced immune suppression. The findings were validated by RNA-Seq, phosphoproteome analysis, and single-cell mass cytometry.

Notably, we observed an upregulation of the *HER2* signalling pathway in the germlines of HER2-negative breast cancer patients, suggesting that *HER2* signalling pathway activity is an important characteristic of HER2-negative breast cancer that can distinguish it from HER2-positive breast cance. As the germline *HER2* signalling pathway was activated in a receptor-independent manner, we presented different models for the activation of the *HER2* signalling pathway and its contribution to the pathogenesis of the two breast cancer subtypes (Suppl. Fig. 4). In HER2-positive breast cancer patients, the *HER2* signalling pathway is activated in tumour tissue due to the amplification of the *HER2* genes [31-33], but remains unchanged in the germlines. Meanwhile, in HER2-negative breast cancer patients, the *HER2* signalling pathway is upregulated by the germline mutations, but there is no amplification of *HER2* genes in the tumour tissue. According to a previous study, the upregulation of *HER2* signalling pathway may lead to immune suppression via *AKT1-*mediated disruption of *STING* signalling [4], which supports our DAGM findings.

Although the APSP risk score from DAGM and PRS both provide disease risk assessment based on the DNA mutations in the germline genome, they are different in the following respects (Suppl. Table 2). First, DAGM uses rare coding variants (minor allele frequency, MAF<1%) from whole-exome sequencing to derive the APSP risk score, whereas PRS generally uses common variants (MAF>5%) from large genome-wide association studies (GWAS), of which nearly 90% lie within non-coding regions of the genome [34]. As GWAS results are ancestry-dependent, it is difficult for PRS to be applied across populations, whereas DAGM was validated using data from a different population (Suppl. Fig. 3). Second, DAGM projects DNA mutations and gene expression onto signalling pathway activities, whereas PRS assesses only SNPs. As a result, DAGM could reveal not only genetic mutations, but also functional signalling pathways, which are biologically relevant to disease. In the future, DAGM could be improved by adding more cell lines and signalling pathways data. Third, PRS is suitable for the risk assessment of complex diseases such as breast cancer with an AUC ranging from 0.63 to 0.69 [9,12], whereas DAGM was used to assess breast cancer risk with an AUC ranging from 0.74 to 0.79. A broader investigation of the application of DAGM in different cancers besides breast cancer is currently underway.

Overall, the DAGM framework provides an effective risk assessment for HER2-negative breast cancer using information encoded in the germline genome, which can facilitate screening of those at high risk of HER2-negative breast cancer for primary prevention. The results from DAGM also provide new insight into the pathogenesis mechanism of breast cancer.

## Abbreviations

APSP: activity profiles of signaling pathway
AUC: area under the receiver operating characteristic curve
CCLE: Cancer Cell Line Encyclopedia
DAGM: damage assessment framework of genomic mutations
ER: Estrogen receptor
GDF: global driving force
GSEA: Gene Set Enrichment Analysis
GWAS: genome-wide association study
HER2: human epidermal growth factor 2
PBMC: peripheral blood mononuclear cells
PCC: Pearson correlation coefficients
PR: Progesterone receptor
PRS: polygenic risk score
RCM: rare coding mutations
SVM: Support Vector Machine
TCGA: The Cancer Genome Atlas
TNBC: Triple-negative breast cancer
WES: whole-exome sequencing.

## Acknowledgements

The authors thank Yanxiang Ni for the comments and suggestions on the manuscript.

## Author contributions

GN and YF conceived the experiments. GN, KW, MY, YF, and ZYW designed the experiments. MY, XLL, TZ, MC, JX, CY, MH, FJ, LZ, WL, JL, CL, ZL, and HG performed the clinical experiments. YF, GN, QZ, JG, SH, CZ, ZZ, XL, HC, GT, YX, YQS, and MQZ analysed the data. GN and YF developed the methodology. YF, MY, ZF, KW, and GN wrote the paper. All authors discussed the results and contributed to the final manuscript.

## Figure Legends

**Suppl. Fig. 1.**
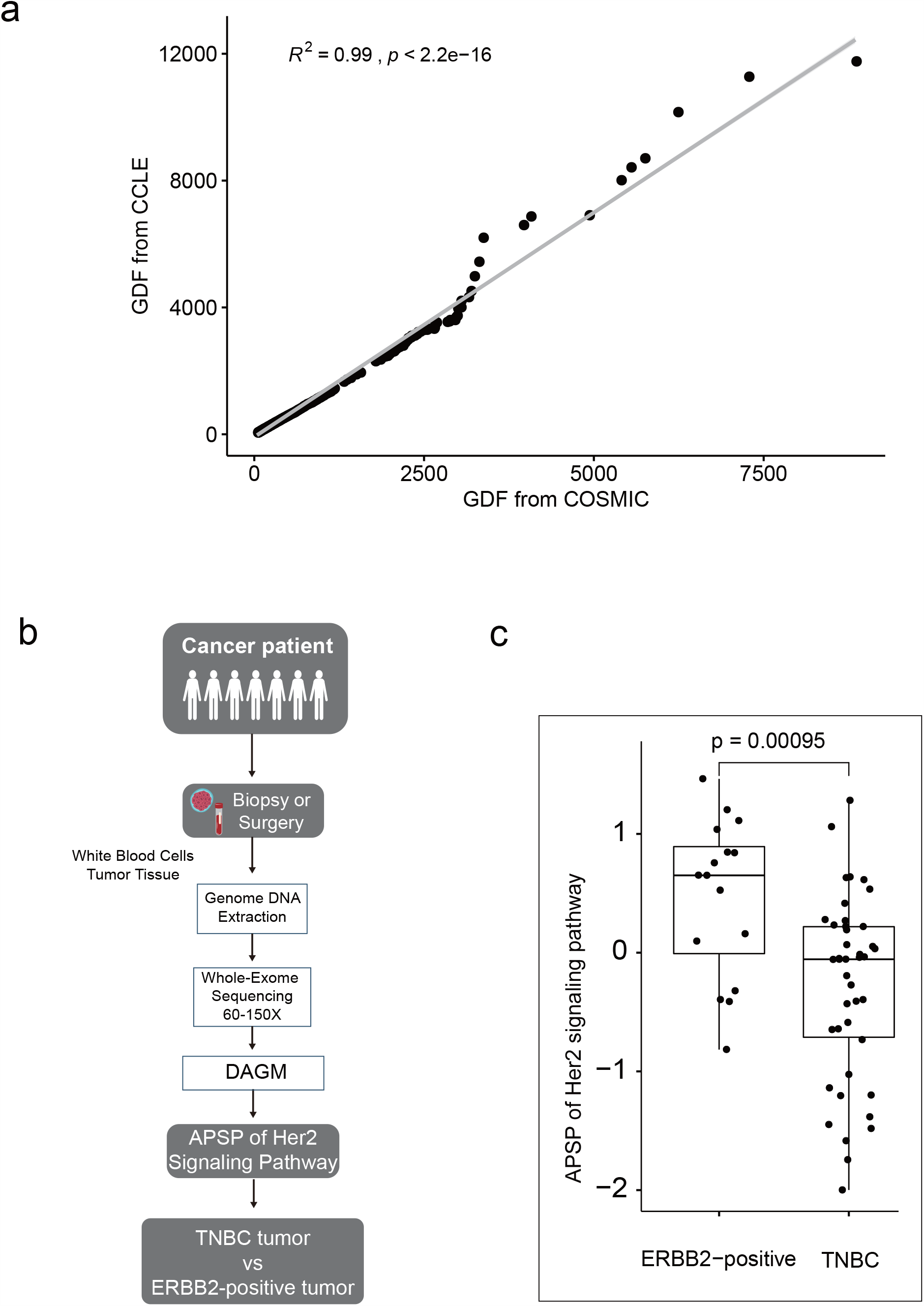
Validations of DAGM framework. (a) Scatter plot of the global driving forces calculated from two different datasets, CCLE and COSMIC. (b and c) Validating DAGM framework in predicting activity of HER2 signalling pathway. The workflow reveals that somatic mutations of breast cancer patients were subjected to DAGM framework to obtain ASPS of Her2 signalling pathway for each subject (b). Of note, the somatic mutations used here does not include information of HER gene amplification. Significant higher APSPs of HER2 signalling pathway were found in ERBB2-positive breast tumour tissues compared to TNBC tissues (c), which is consistent with the pathological diagnosis by IHC and HER2-FISH.

**Suppl. Fig. 2.**
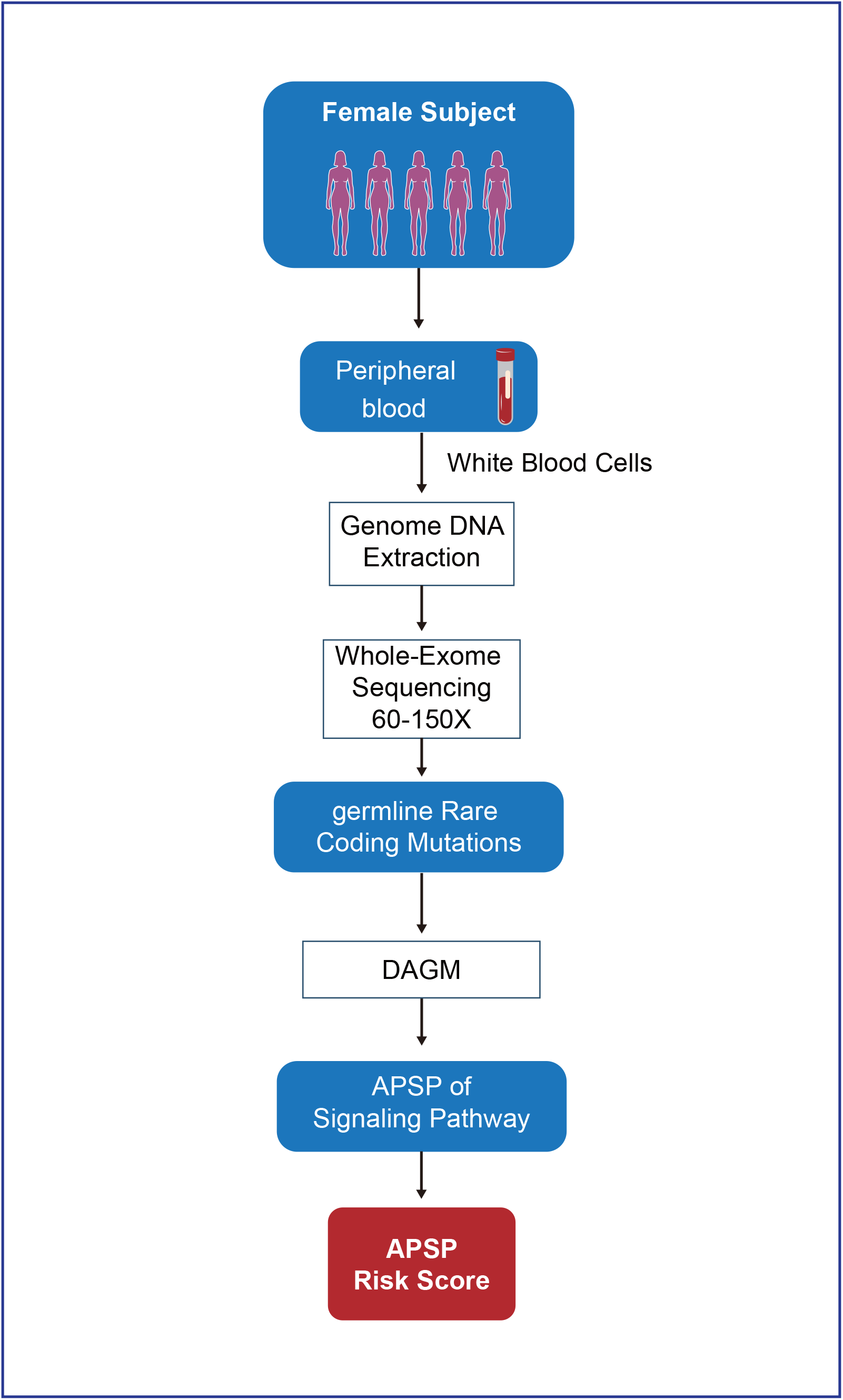
Workflow of DAGM using germline mutations to calculate the activity profile of signalling pathways.

**Suppl. Fig. 3.**
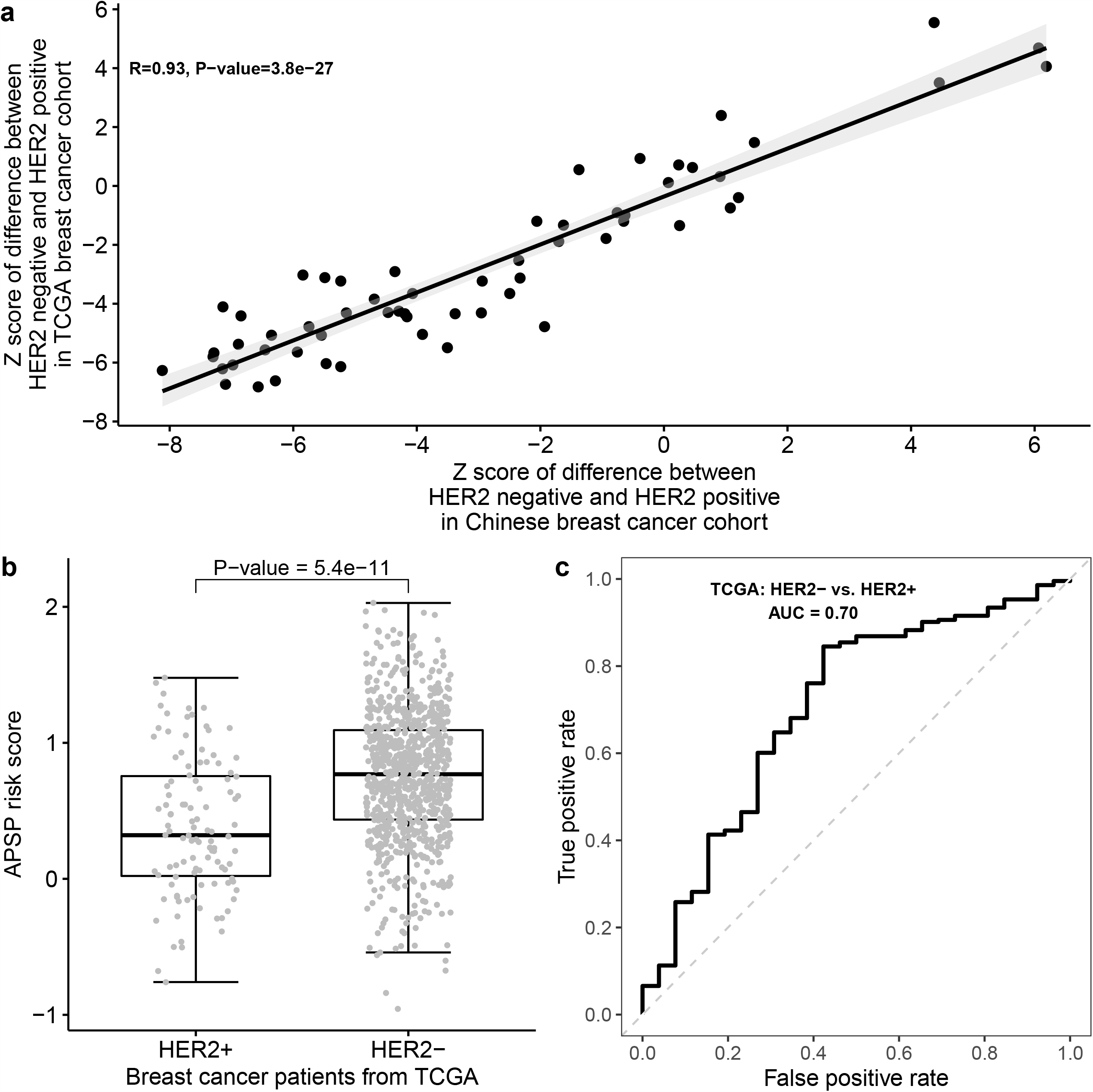
Validation using breast cancer patient data available in The Cancer Genome Atlas (TCGA). (a) Z-score of activity difference between HER2-positive and HER2-negative patients was obtained for each signalling pathway in Chinese or TCGA cohorts. Scatter plot shows significant correlation of these Z-scores between Chinese and TCGA cohorts. (b) Boxplot of APSP risk scores of HER2-negative or HER2-positive breast cancer patients in TCGA. (c) Receiver operating characteristic curve of the classifier between HER2-negative and HER2-positive breast cancer patients in TCGA based on the APSP risk scores. Area under the curve (AUC) was 0.70.

**Suppl. Fig. 4.**
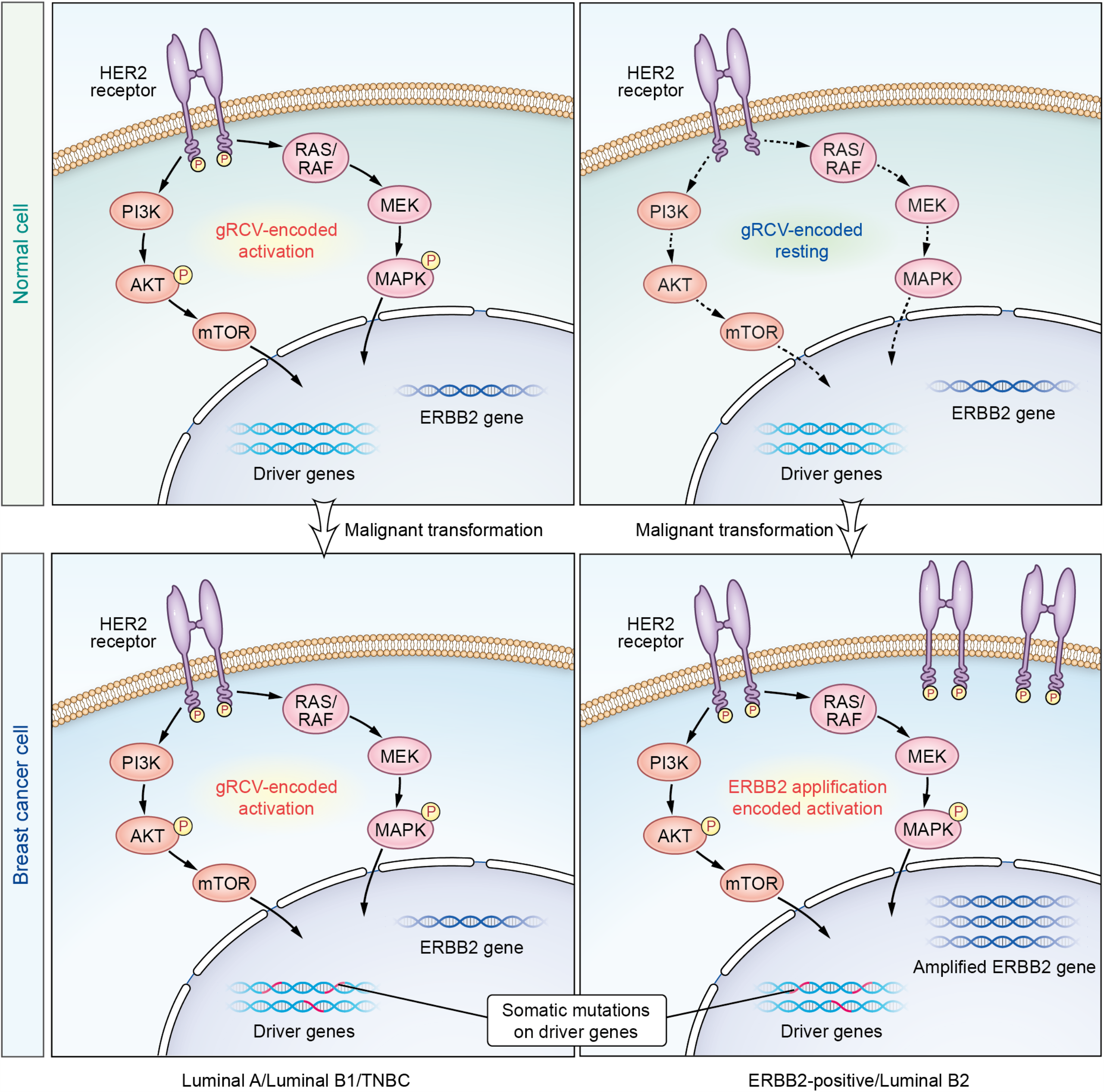
HER2 signalling pathway was activated by germline rare coding mutations in HER2-negative breast cancer. Germline rare coding mutations inferred the activation of HER2 signalling pathway could explain the potential pathogenesis of HER2-negative breast cancer (TNBC, Luminal A, and Luminal B (HER2-)). On the other hand, unchanged HER2 signalling pathway inferred by germline rare coding mutations could explain the pathological type of HER2-positive breast cancer (ERBB2-positive and Luminal B (HER2+)).

**Supplementary Table 1.** Antibodies used for CyTOF.

| Index | Antibody |
| --- | --- |
| 1 | CD45 |
| 2 | CD25 |
| 3 | CD28 |
| 4 | CD66b |
| 5 | CD3 |
| 6 | CD57 |
| 7 | CD152 (CTLA-4) |
| 8 | CD366 (TIM-3) |
| 9 | IgM |
| 10 | CD38 |
| 11 | FoxP3 |
| 12 | CD86 |
| 13 | CD56 |
| 14 | CD39 |
| 15 | CD137 (4-1BB) |
| 16 | CD279 (PD-1) |
| 17 | CD19 |
| 18 | CD161 |
| 19 | CD64 |
| 20 | CD16 |
| 21 | CD196 (CCR6) |
| 22 | CD154 (CD40L) |
| 23 | IFN-γ |
| 24 | HLA-DR |
| 25 | CD14 |
| 26 | CD45RA |
| 27 | TLR2 (CD282) |
| 28 | CD4 |
| 29 | IgD |
| 30 | CD194 (CCR4) |
| 31 | CD45RO |
| 32 | CD8a |
| 33 | CD123 (IL-3Ra) |
| 34 | CD68 |
| 35 | Tbet |
| 36 | CD11b |
| 37 | CD183 (CXCR3) |
| 38 | CD197 (CCR7) |
| 39 | CD185 (CXCR5) |
| 40 | CD33 |
| 41 | CD11c |
| 42 | IL-7Ra (CD127) |

**Suppl Table 2.** Comparison between APSP risk score and polygenic risk score.

| Method | APSP risk score | Polygenic risk score |
| --- | --- | --- |
| Sample size for modeling | Hundreds | Tens of thousands |
| Original data source | WES | GWAS |
| Based on germline genome? | Yes | Yes |
| **Variants frequency** | **Rare (MAF<1%)** | **Common (MAF>5%)** |
| **Variants type** | **Coding** | **Majority of non-coding** |
| Number of applied variants | Thousands | Dozens to thousands |
| Weighted factor | APSP | Variant allele |
| Weights | Constant | Log odds ratio |
| **Supporting biological mechanism?** | **Yes** | **No** |
| **Ancestry specific?** | **No** | **Yes** |
| Generalize to other population? | Yes | No |
| Compatible with BRCA1/2? | Significant | NA |
| **AUC for breast cancer** | 0.74-0.79 | 0.63-0.69 |

## References

[1] Jiang X, Tang H, Chen T. Epidemiology of gynecologic cancers in China. Journal of gynecologic oncology 2018;29:e7.

[2] Siegel RL, Miller KD, Jemal A. Cancer statistics, 2018. CA: a cancer journal for clinicians 2018;68:7–30.

[3] Sun J, Meng H, Yao L, Lv M, Bai J, Zhang J et al. Germline Mutations in Cancer Susceptibility Genes in a Large Series of Unselected Breast Cancer Patients. Clin Cancer Res 2017;23:6113–6119.

[4] Wu S, Zhang Q, Zhang F, Meng F, Liu S, Zhou R et al. HER2 recruits AKT1 to disrupt STING signalling and suppress antiviral defence and antitumour immunity. Nat Cell Biol 2019;21:1027–1040.

[5] Sivick KE, Desbien AL, Glickman LH, Reiner GL, Corrales L, Surh NH et al. Magnitude of Therapeutic STING Activation Determines CD8 T Cell-Mediated Anti-tumor Immunity. Cell reports 2018;25:3074-3085.e3075.

[6] Lei J, Rudolph A, Moysich KB, Rafiq S, Behrens S, Goode EL et al. Assessment of variation in immunosuppressive pathway genes reveals TGFBR2 to be associated with prognosis of estrogen receptor-negative breast cancer after chemotherapy. Breast cancer research : BCR 2015;17:18.

[7] Subramanian A, Tamayo P, Mootha VK, Mukherjee S, Ebert BL, Gillette MA et al. Gene set enrichment analysis: A knowledge-based approach for interpreting genome-wide expression profiles. PNAS 2005;102:15545–15550.

[8] Sugrue LP, Desikan RS. What Are Polygenic Scores and Why Are They Important? JAMA 2019;321:1820–1821.

[9] Mavaddat N, Michailidou K, Dennis J, Lush M, Fachal L, Lee A et al. Polygenic Risk Scores for Prediction of Breast Cancer and Breast Cancer Subtypes. Am J Hum Genet 2019;104:21–34.

[10] Li H, Feng B, Miron A, Chen X, Beesley J, Bimeh E et al. Breast cancer risk prediction using a polygenic risk score in the familial setting: a prospective study from the Breast Cancer Family Registry and kConFab. Genet Med 2017;19:30–35.

[11] Läll K, Lepamets M, Palover M, Esko T, Metspalu A, Tõnisson N et al. Polygenic prediction of breast cancer: comparison of genetic predictors and implications for risk stratification. BMC cancer 2019;19:557.

[12] Khera AV, Chaffin M, Aragam KG, Haas ME, Roselli C, Choi SH et al. Genome-wide polygenic scores for common diseases identify individuals with risk equivalent to monogenic mutations. Nature genetics 2018;50:1219–1224.

[13] Wang B, Bao S, Zhang Z, Zhou X, Wang J, Fan Y et al. A rare variant in MLKL confers susceptibility to ApoE l’.4-negative Alzheimer’s disease in Hong Kong Chinese population. Neurobiol Aging 2018;68:160.e161-160.e167.

[14] Li H. Aligning sequence reads, clone sequences and assembly contigs with BWA-MEM. q-bio.GN 2013; 1303.3997v2:

[15] McKenna AH, Hanna M, Banks E, Sivachenko A, Cibulskis K, Kernytsky A et al. The Genome Analysis Toolkit: A MapReduce framework for analyzing next-generation DNA sequencing data. Genome Res 2010;20:1297–1303.

[16] Wang K, Li M, Hakonarson H. ANNOVAR: functional annotation of genetic variants from high-throughput sequencing data. Nucleic Acids Res 2010;38:e164–e164.

[17] Kim D, Paggi JM, Park C, Bennett C, Salzberg SL. Graph-based genome alignment and genotyping with HISAT2 and HISAT-genotype. Nature Biotechnology 2019;37:907–915.

[18] Liao Y, Smyth GK, Shi W. featureCounts: an efficient general purpose program for assigning sequence reads to genomic features. Bioinformatics 2014;30:923–930.

[19] Robinson MD, McCarthy DJ, Smyth GK. edgeR: a Bioconductor package for differential expression analysis of digital gene expression data. Bioinformatics 2010;26:139–140.

[20] Consortium TU. UniProt: a worldwide hub of protein knowledge. Nucleic Acids Research 2018;47:D506–D515.

[21] Murtagh F, Legendre P. Ward’s Hierarchical Agglomerative Clustering Method: Which Algorithms Implement Ward’s Criterion? J Classif 2014;31:274–295.

[22] Ghandi M, Huang FW, Jané-Valbuena J, Kryukov GV, Lo CC, McDonald ER et al. Next-generation characterization of the Cancer Cell Line Encyclopedia. Nature 2019;569:503– 508.

[23] Sondka Z, Bamford S, Cole CG, Ward SA, Dunham I, Forbes SA. The COSMIC Cancer Gene Census: describing genetic dysfunction across all human cancers. Nature Reviews Cancer 2018;18:696–705.

[24] Liu S-H, Shen P-C, Chen C-Y, Hsu A-N, Cho Y-C, Lai Y-L et al. DriverDBv3: a multi-omics database for cancer driver gene research. Nucleic Acids Research 2019;

[25] Dai X, Li T, Bai Z, Yang Y, Liu X, Zhan J et al. Breast cancer intrinsic subtype classification, clinical use and future trends. American journal of cancer research 2015;5:2929–2943.

[26] Antoniou A, Pharoah PDP, Narod S, Risch HA, Eyfjord JE, Hopper JL et al. Average risks of breast and ovarian cancer associated with BRCA1 or BRCA2 mutations detected in case Series unselected for family history: a combined analysis of 22 studies. Am J Hum Genet 2003;72:1117–1130.

[27] Wong-Brown MW, Meldrum CJ, Carpenter JE, Clarke CL, Narod SA, Jakubowska A et al. Prevalence of BRCA1 and BRCA2 germline mutations in patients with triple-negative breast cancer. Breast cancer research and treatment 2015;150:71–80.

[28] Levy-Lahad E, Friedman E. Cancer risks among BRCA1 and BRCA2 mutation carriers. British journal of cancer 2007;96:11–15.

[29] Shieh Y, Fejerman L, Lott PC, Marker K, Sawyer SD, Hu D et al. A Polygenic Risk Score for Breast Cancer in US Latinas and Latin American Women. JNCI: Journal of the National Cancer Institute 2019;112:590–598.

[30] Bauer S, Robinson PN, Gagneur J. Model-based gene set analysis for Bioconductor. Bioinformatics 2011;27:1882–1883.

[31] Tai W, Mahato R, Cheng K. The role of HER2 in cancer therapy and targeted drug delivery. J Control Release 2010;146:264–275.

[32] Andrechek ER. HER2/Neu tumorigenesis and metastasis is regulated by E2F activator transcription factors. Oncogene 2015;34:217–225.

[33] Zaczek A, Brandt B, Bielawski KP. The diverse signaling network of EGFR, HER2, HER3 and HER4 tyrosine kinase receptors and the consequences for therapeutic approaches. Histol Histopathol 2005;20:1005–1015.

[34] Giral H, Landmesser U, Kratzer A. Into the Wild: GWAS Exploration of Non-coding RNAs. Frontiers in Cardiovascular Medicine 2018;5:

